# Molecular diagnosis of SARS-CoV-2: a validation of saliva samples

**DOI:** 10.1101/2022.01.20.22269618

**Authors:** Natalia Verissimo Rodrigues da Silva, Samyra Stéphane Neves Pereira, Karine Frehner Kavalco, Fabiano Bezerra Menegídio, Luanda Medeiros-Santana, Liliane Evangelista Visôtto, Rubens Pasa

**Affiliations:** Laboratory of Molecular Diagnosis, Federal University of Viçosa, campus Rio Paranaíba. Road MG 230 Km 7, LAE Building, room 237, Zip code 38810-000, Rio Paranaíba – MG, Brazil

**Keywords:** SARS-CoV-2, RT-qPCR, Covid-19, nasopharyngeal swabs (NPS)

## Abstract

Nasopharyngeal swabs are the most used in sample collecting for covid-19 tests in SARS-CoV-2 molecular diagnosis. However, this sampling method presents some disadvantages, since, in addition to being dependent on imported materials, it is invasive, causes discomfort in patients, and, presents the risk of contamination for the medical collection team. This study aimed at validating saliva samples to obtain viral RNA to be used in the molecular diagnostic test for SARS-CoV-2 using the RT-qPCR technique. Results presented 93,44% concordance in in comparison to nasopharyngeal swabs sampling. Therefore, saliva samples used in SARS-CoV-2 RT-qPCR detection tests presented consistent results.

## INTRODUCTION

The protein Spike (S), an antigen found on the surface of SARS-CoV-2, is one of the main means of infection of the virus. It binds to the receptor ACE2, an angiotensin-converting enzyme, found in several epithelial cells of the respiratory tract and salivary gland ducts of the host (DIAS et al., 2020).

The infection of SARS-CoV in the salivary glands of monkeys showed a high affinity between the viral antigen and the ACE2 protein of the host (LIU et al., 2011). This way, the presence of the virus SARS-CoV-2 in the human saliva reinforces the possibility of salivary glandule infection (TO et al 2020).

Collection of samples for covid-19 tests through the nasopharynges for swabs are the most used in RT-qPCR molecular diagnostic tests. However, this collection method is not only invasive and painful for the patient, being counter-indicated in some cases, but it also requires some training from the medical staff and put health professionals at risk of contamination due to their proximity to patients and the consequent exposure to the virus (TO et al 2020).

Saliva samples, in turn, can be easily collected by the patient without the need for invasive procedures, bringing other benefits, as the decrease in the exposition of health professionals, no counter-indications, and no need for a viral transport solution, just a sterile container (ECHAVARRIA et al 2021; TO et al 2020).

This study aimed at validating the use of saliva from patients as a sample for Covid-19 molecular diagnostic tests in the Alto Paranaíba region, Minas Gerais, Brazil.

## MATERIAL AND METHODS

Between the months of August and September 2020, 61 paired samples, nasopharyngeal swabs (NPS) and saliva, were collected from asymptomatic and symptomatic patients with suspected SARS-CoV-2 contamination, submitted by the Regional Health Superintendence of *Patos de Minas, Minas Gerais*, Brazil. The samples were collected from patients presenting COVID-19 symptoms, such as fever, sore throat, headache, cough, absence of smell and taste, and from asymptomatic patients who were in contact with either positive or suspect cases.

Health professionals were responsible for collecting the NPS samples, conditioned in a saline solution, while patients were guided by them during the self-collection of their saliva samples in sterile containers. All samples were kept in a cooler at 4°C, maximum, until the Laboratory of Molecular Diagnosis of Federal University of Viçosa, campus Rio Parnaíba, Minas Gerais State, where the molecular analysis were conducted.

The samples were first vortexed in a biosafety cabinet NB2. RNA extraction was conducted using 200 μl of the samples following the manual column extraction with the Bio Gene Kit from Bioclin.. After RNA extraction RT-qPCR assay was carried out using the Allplex™ 2019-nCov Assay (Seegene) kit, which identifies three SARS-CoV-2 target genes (RdRP, N, and E) (Fig. 1). Following the manufacturer’s instruction, internal control was added before RNA isolation. The protocol was composed of 3 cycles: i) 20 minutes at 50°C; ii) 15 minutes at 95°C; and iii) 45 cycles of 15 seconds at 94°C and 30 seconds at 58°C. The assays were conducted on the CFX-96 Real-Time Cycler (Bio-Rad). Purified water was used as internal negative control and sequences of amplification genes for positive control, as instructed by the manufacturer.

**Figure 1.**
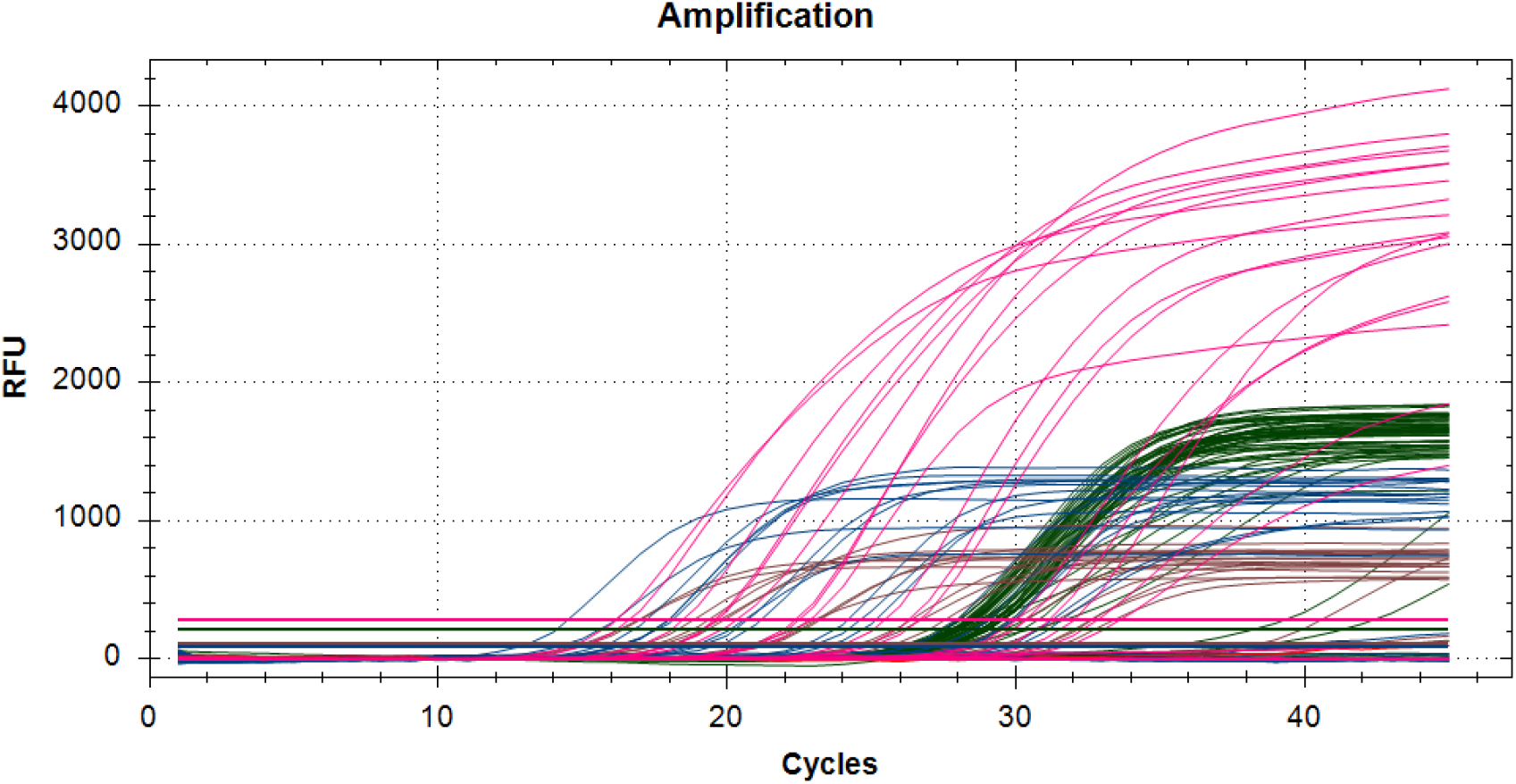
RT-qPCR amplification of SARS-CoV-2 detection. Each color represents the dynamic of amplification cycles of the viral genes RdRP (in pink), E (in red) and N (in blue), and internal control (in green).

Positive results were considered when at least one of the target genes was amplified before the 40th cycle of the RT-qPCR during the cycle threshold (Ct), regardless of the internal amplification control.

## RESULTS AND DISCUSSION

Results presented 93,44% concordance in both sampling methods. Among the results of paired samples that were completely concordant, 15 were positive while 42 were not. The virus was detectable exclusively through nasopharyngeal swab in only one sample, and in other three samples, it was only detected on saliva (Fig. 2).

**Figure 2.**
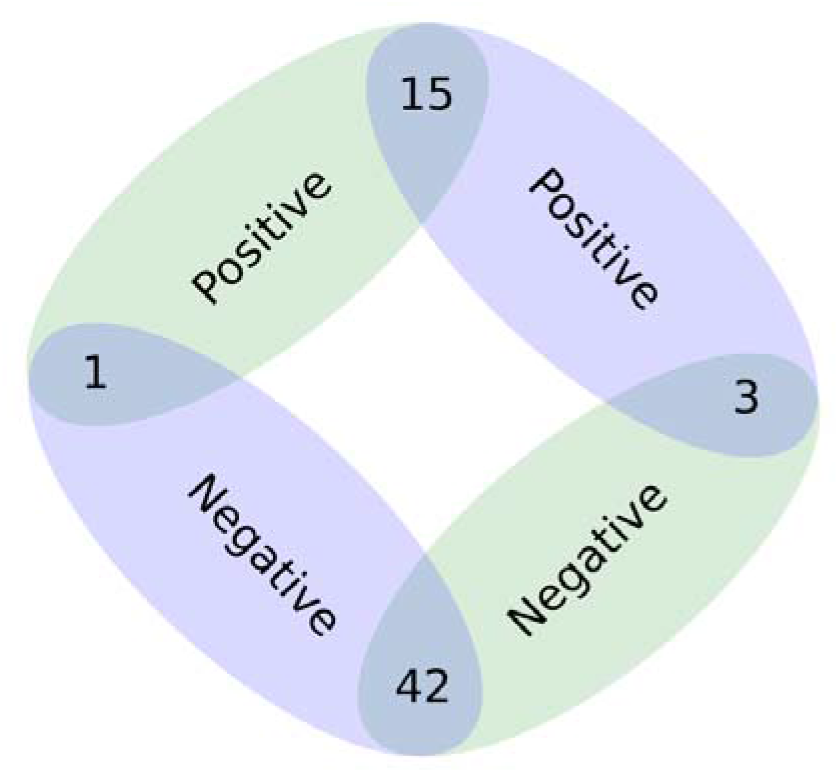
Venn diagram illustrating the concordance among obtained results. Nasopharyngeal samples in green and saliva samples in blue.

Studies have shown a high detection rate in saliva samples from asymptomatic (RAO et al., 2020) and symptomatic patients (PASOMSUB et al., 2020) by RT-qPCR. In both studies, SARS-CoV-2 infections, which were not detected in nasopharyngeal swab samples, were detected in saliva, suggesting they can have greater detection sensitivity as the tongue and salivary glands are possibly the main sites of infection, replication, and both direct and indirect transmission of SARS-CoV-2 (TO et al., 2020; XU et al., 2020).

Molecular diagnostics of SARS-CoV-2 via RT-qPCR using saliva samples is proving to be an alternative to nasopharyngeal swab collection methods.

## Data Availability

All data produced in the present study are available upon reasonable request to the authors

## ETHICAL APPROVAL

The university’s institutional review board approved the analysis and issued a waiver of consent (Ethics Committee Ruling number – CAAE: 33446820.8.0000.5153).

## ACKNOWLEDGEMENTS

The authors would like to thank all the team at Laboratory of Molecular Diagnosis of UFV, campus Rio Paranaíba, for the support when conducting the diagnostic tests.

